# Automated Extraction of Mortality Information from Publicly Available Sources Using Language Models

**DOI:** 10.1101/2024.10.28.24316027

**Authors:** Mohammed Al-Garadi, Michele LeNoue-Newton, Michael E. Matheny, Melissa McPheeters, Jill M. Whitaker, Jessica A. Deere, Michael F. McLemore, Dax Westerman, Mirza S. Khan, José J. Hernández-Muñoz, Xi Wang, Aida Kuzucan, Rishi J. Desai, Ruth Reeves

**Affiliations:** Department of Biomedical Informatics, Vanderbilt University Medical Center, Nashville, TN, USA; Geriatrics Research Education and Clinical Care Service, Tennessee Valley Healthcare System VA, Nashville, TN, USA; RTI International, Research Triangle Park, NC, US; Office of Surveillance and Epidemiology, Center for Drug Evaluation and Research, US Food and Drug Administration, Silver Spring, MD; Division of Pharmacoepidemiology and Pharmacoeconomics, Brigham and Women’s Hospital and Harvard Medical School, Boston, MA, USA

## Abstract

**Background:** Mortality is a critical variable in healthcare research, especially for evaluating medical product safety and effectiveness. However, inconsistencies in the availability and timeliness of death date and cause of death (CoD) information present significant challenges. Conventional sources such as the National Death Index (NDI) and electronic health records (EHRs) often suffer from data lags, missing fields, or incomplete coverage, limiting their utility in time-sensitive or large-scale studies. With the growing use of social media, crowdfunding platforms, and online memorials, publicly available digital content has emerged as a potential supplementary source for mortality surveillance. Despite this potential, accurate tools for extracting mortality information from such unstructured data sources remain underdeveloped.

**Objective:** To develop scalable approaches using natural language processing (NLP) and large language models (LLM) for the extraction of mortality information from publicly available online data sources, including social media platforms, crowdfunding websites, and online obituaries, and to evaluate their performance across various sources.

**Methods:** Data were collected from public posts on X (formerly Twitter), GoFundMe campaigns, memorial websites (EverLoved.com and TributeArchive.com), and online obituaries from 2015 to 2022, focusing on U.S.-based content relevant to mortality. We developed an NLP pipeline using transformer-based models to extract key mortality information such as decedent names, dates of birth, and dates of death. We then employed a few-shot learning (FSL) approach with LLMs to identify primary and secondary causes of death. Model performance was assessed using precision, recall, F1-score, and accuracy metrics, with human-annotated labels serving as the reference standard for the transformer-based model and a human adjudicator blinded to labeling source for the FSL model reference standard.

**Results:** The best-performing model obtained a micro-averaged F1-score of 0.88 (95% CI, 0.86-0.90) in extracting mortality information. The FSL-LLM approach demonstrated high accuracy in identifying primary CoD across various online sources. For GoFundMe, the FSL-LLM achieved 95.9% accuracy for primary cause identification, compared to 97.9% for human annotators. In obituaries, FSL-LLM accuracy was 96.5% for primary causes, while human accuracy was 99.0%. For memorial websites, FSL-LLM achieved 98.0% accuracy for primary causes, with human accuracy at 99.5%.

**Conclusions:** This study demonstrates the feasibility of using advanced NLP and LLM techniques to extract mortality data from publicly available online sources. These methods can significantly enhance the timeliness, completeness, and granularity of mortality surveillance, offering a valuable complement to traditional data systems. By enabling earlier detection of mortality signals and improving CoD classification across large populations, this approach may support more responsive public health monitoring and medical product safety assessments. Further work is needed to validate these findings in real-world healthcare settings and facilitate the integration of digital data sources into national public health surveillance systems.

## Introduction

Mortality is a critical variable in healthcare research, and all-cause mortality is one of the most studied endpoints[1–4]. Accurate identification of the fact, timing, and cause of death (CoD) is essential for various types of medical research, including clinical trials, observational studies, and post-marketing surveillance programs such as the US FDA Sentinel System [5–8].

A recent report identified limitations in the availability of date and CoD information as a major cause for study insufficiency when considering the use of the Sentinel Active Risk Identification and Analysis (ARIA) system to address regulatory questions[9]. Failing to identify deaths may result in substantial underestimation of mortality outcomes related to medical products, so efforts to identify additional data sources to supplement current systems have far-reaching consequences. Vital statistics data, collected in the U.S. through death certificates and submitted at the state level, serve as the ‘reference standard’ for mortality information. Depending on U.S. state laws (and sometimes the manner of death), death certificates may be completed by coroners, medical examiners, or physicians within the healthcare system. Once submitted to state systems, death certificate data are forwarded to the Centers for Disease Control and Prevention (CDC), which codes the underlying CoD and adds it to national records. However, this process is slow—vital statistics data are typically delayed by at least nine months, and the National Death Index (NDI) often lags by up to two years.There are other data sources for death information, including claims databases, and medical records, but each of these sources has limitations[10, 11]. Claims databases may underrepresent uninsured populations, while medical records often lack standardization between healthcare providers, complicating data aggregation and comparison[12]. In most claims databases, death-related information, including occurrence and CoD, is often incomplete or not directly recorded. Similarly, healthcare system-based data sources, such as electronic health records (EHRs), frequently lack comprehensive mortality data, particularly when patients are not under the care of the healthcare system at the time of death. This poses significant challenges for researchers and clinicians relying on these data sources for epidemiological studies, outcomes research, and healthcare quality assessments.

The rise in the use of social media has introduced potential sources of mortality-related information, including online obituaries and the sharing of death information in social networks through Twitter and other channels. There is growing precedent for the use of social media in public health and other health-related research, and user posts have been used to track illnesses[13–17], measure behavioral risk factors[18–22], localize diseases geographically[21, 23, 24], and analyze symptoms and medication usage[25–30]. Nonetheless, a key challenge inherent in social media data for mortality information is the capacity to extract the data and CoD at scale and with replicable methods. These social media sources offer potential advantages in timeliness, context, and coverage compared to traditional mortality data sources.

In this study, we sought to develop a set of NLP tools to extract both the fact and CoD from publicly available records and to assess the relative information density of illness and death information within these records. This type of data, when combined with other sources, could improve ascertainment in downstream studies that require the use of the facts and causes of mortality among EHR and claims data analyses.

The innovative approach leverages publicly available data to provide timely insights into population health trends, potentially enabling faster responses to emerging health threats. By linking social media and obituary data with patient records, the system could offer a more comprehensive view of health outcomes and risk factors, as well as system evaluation.

This study highlights the transformative potential of LLMs in comparison to traditional NLP approaches. While earlier methods rely heavily on predefined rules or extensive labeled datasets, LLMs offer greater flexibility through few-shot learning and contextual understanding of complex narratives[31]. This capability is especially critical for extracting nuanced mortality information from diverse and informal online text, where structure and terminology often vary widely across sources.

In this study, we developed and evaluated a pipeline combining transformer-based NLP models and few-shot learning with LLMs to extract mortality information, including fact and cause of death, from publicly available online sources. Our goal was to assess the feasibility, accuracy, and utility of this approach for supplementing traditional mortality data in healthcare research and surveillance.

## Methods

We developed and evaluated natural language processing (NLP) techniques to extract mortality information from publicly available online sources, focusing on U.S.-related data. The research methodology included data collection, NLP model development, and performance assessment. Given its focus on public health surveillance using open-source information, this research qualifies for exemption from FDA and Vanderbilt University Medical Center (VUMC) Institutional Review Board (IRB) oversight.

### Data Sources and Study Cohort

Data were collected from X (formerly Twitter), GoFundMe, and online obituaries (Obituaries.com), memorial websites (EverLoved.com, and TributeArchive.com) between the years 2015 and 2022 in the United States, which are publicly available and aggregated for research purposes in accordance with fair use. The online obituary sources provide more robust meta-data for determining inclusion criteria than records obtained from Twitter or GoFundMe. Our collection methods, therefore, differed by source.

Our search on X (formerly Twitter) utilized around 50 derived keywords (the list is in the supplementary material) for English-language posts while excluding non-English content. Keywords included terms like ‘death,’ ‘expired,’ and ‘deceased.’ Using Twitter’s official research Application Programming Interface (API), this approach yielded approximately 40 million tweets. Using similar keywords (provided in the appendix), we identified and retrieved posts from GoFundMe and memorial websites (EverLoved.com and TributeArchive.com) containing mortality-related information. For Obituaries.com, we acquired reports from 2015 to 2022, which contained millions of records (see Table 5 for final counts across all sources). For the obituary data sources, we collected structured metadata (e.g., first name, last name, date of death, date of birth, location) and extracted accompanying textual information. NLP techniques were subsequently used on this textual content to supplement or complete missing or incomplete metadata fields. This approach allowed us to maximize the information extracted from each obituary, enhancing the overall quality and completeness of our dataset.

### Reference Standard

To construct a human-based reference dataset for training and testing our models, we developed an annotation process that captured the deceased’s name, names of related individuals, key dates (including death, birth, and other relevant dates), and CoD. First, annotators were instructed to accurately classify names with post-nominals, avoid names in Twitter handles, and use specific relationship attributes for related persons (e.g., spouse, sibling, child). Second, annotated dates included exact, partial, or relative expressions, with clear distinctions for death and birth dates. Third, the causes of death were annotated with attributes indicating assertion (positive, negative, uncertain) and patient vs. not-patient (reference to the deceased or to someone else). Finally, if no relevant data was found in a document, annotators classified the document as “No Data”.

A corpus of 4,200 notes, 1,050 from each of the data sources, was randomly sampled. We split the 4,200 annotated posts from all data sources into training (70%), testing (20%), and validation (10%) datasets. The training data contained 81,082 tokens (words), and the test data contained 27,834 tokens (words).

### Annotation

The data were annotated by three trained nurse annotators who closely followed a detailed annotation guideline, categorizing each post into first and last names, dates of birth, dates of death, and CoD. The training was initiated using records from Twitter, GoFundMe, the memorial website (EverLoved/TributeArchives), and Obituaries.com, with all three annotators independently labeling the same documents in rounds of 15 documents from each source (n=45). After each training round of annotation was completed by all three members, agreement rates were computed between pairs of annotation sets. The overall inter-annotator agreement (IAA) was evaluated using Cohen’s kappa[32], and annotators were required to achieve an overall IAA threshold of 0.80 on the training set before proceeding with the full annotation process. When the targeted threshold was not met, the annotation team performed a consensus annotation over each document in a given annotation round, discussing their differences, and updating or clarifying the annotation guidelines. Once trained, each annotator independently labeled a subset of a corpus totaling 4,200 documents (1,050 per source). To assess reliability, 100 additional documents (25 per source) were randomly assigned to all three annotators, and an independent IAA was conducted. The eHOST annotation tool was used to annotate the documents.[33]

### Information density assessment

Annotations completed by nurse annotators were used to assess the information density of online sources, such as social media platforms like Twitter, to determine if they contained sufficient details for reliable patient linkage and augmentation of date of death in healthcare systems. Sources with inadequate information were excluded from further analysis.

Assessment of CoD availability was completed using the 600 document annotations utilized in the few-shot learning validation with verification by the nurse adjudicator of causes of death mentioned within the post.

### NLP Development and Implementation

We developed in parallel two NLP tools for information extraction from the previously described social media sources. First, we adapted four deep learning transformer-based methods including BERT (Bidirectional Encoder Representations from Transformers)[34], RoBERTa (Robustly Optimized BERT Pretraining Approach)[35], ALBERT (A Lite BERT)[36], BERTweet[37] to extract the decedent’s name, date of birth, and date of death, and to exclude any irrelevant dates. The technical pipeline overview for the transformer-based model is illustrated in Figure 1.

**Figure 1:**
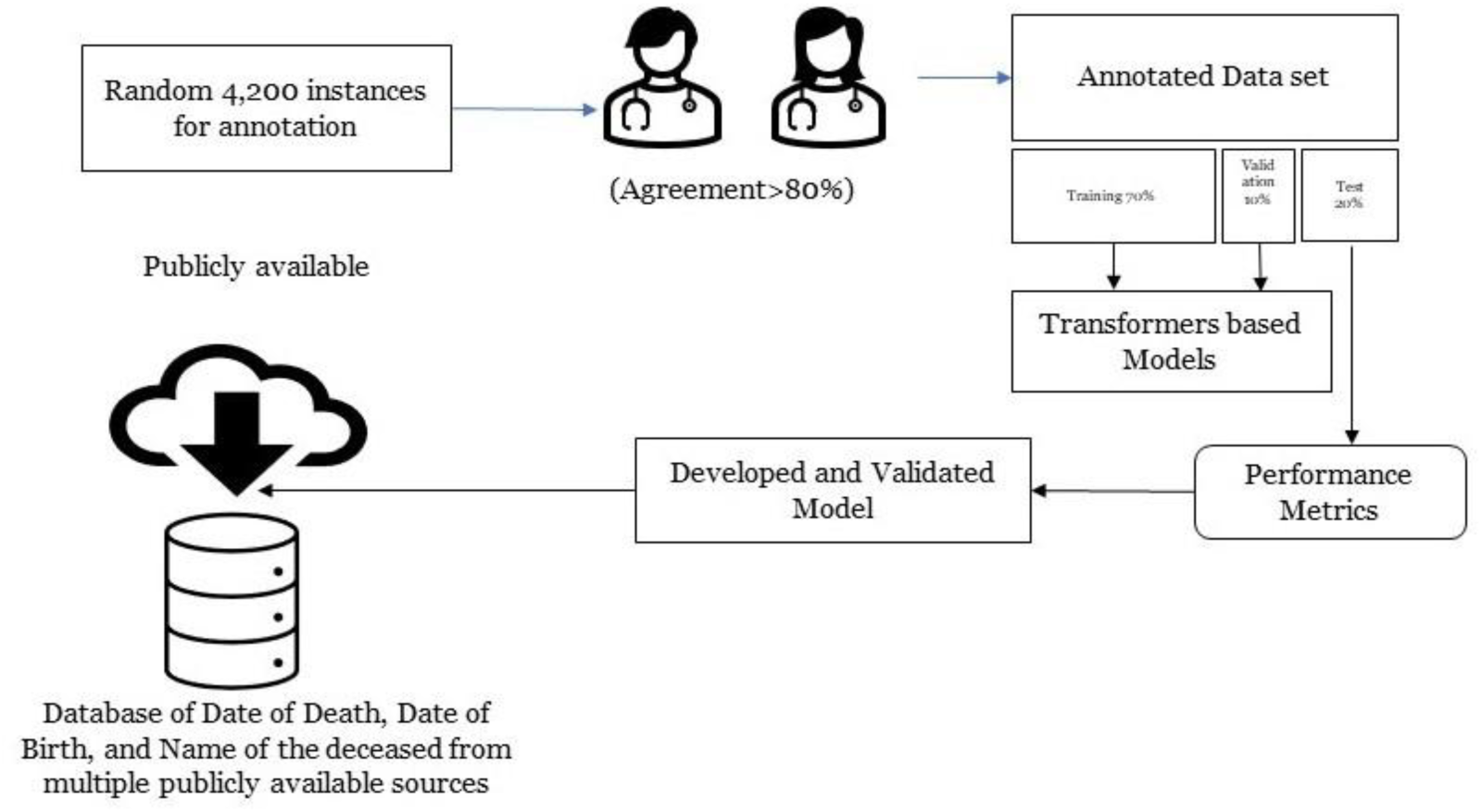
Workflow of the NLP pipeline development and evaluation process.

To identify CoD, we used a few-shot learning approach to leverage an open-source Large Language Model (LLM) (Figure 2). The decision to forgo transformer models for this phase of information extraction was based on the need for a nuanced understanding of both the extracted cause and its contextual relevance in predicting CoD. Instead, we employed an iterative prompting strategy incorporating annotated examples and structured guidelines to delineate primary and secondary CoD. High-quality annotation labels, determined by consensus between at least two annotators, ensured the reliability of the prompts.

**Figure 2:**
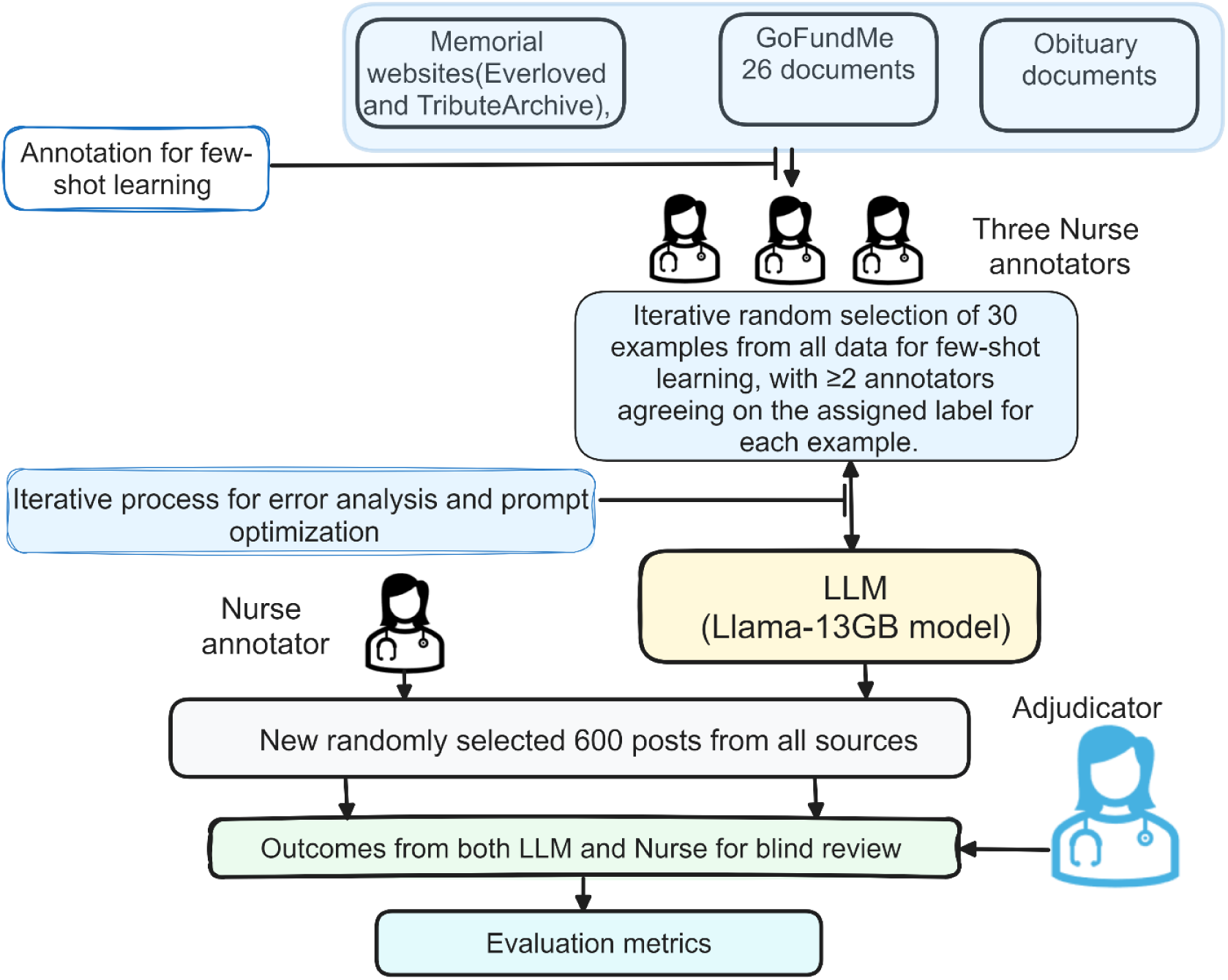
Workflow for Few-Shot Learning and Evaluation.

For example, in the post, “Jane Smith died from a severe infection following surgery. She also had diabetes and hypertension, which contributed to her deteriorating health,” the main cause would be noted as “Severe Infection Following Surgery,” and the secondary causes as “Diabetes” and “Hypertension.” The initial prompt engineering stage ensures the LLM properly formulates the type of information to extract/predict. We utilized the LLaMA model, a 13 GB language model developed by Facebook AI Research, for processing the data[38]. LLaMA, which stands for “Large Language Model Meta AI,” is a foundational language model that exhibits remarkable performance across various NLP tasks[38]. A smaller version, such as the 13GB variant, of the Llama model can be run locally on a machine with sufficient computational resources, making it more accessible and efficient for certain applications. We started with 30 randomly selected examples from the manually annotated data (training split) for prompting (The LLM-Prompt example is available in the supplementary material), and 30 for assessing the model’s performance, where at least two annotators agreed on the annotated instance. The prompts and assessment examples went through several iterations of LLM refinement, totaling four iterations, until the identified CoD was correct in most cases across the various assessment sets. The accuracy during the prompting process was evaluated qualitatively to understand where the model performed correctly and where it made errors.

During the testing phase, we evaluated our final prompting design on a new set of 600 examples. The evaluation process involved three steps:

- A nurse annotator identified the CoD in these examples following the provided guidelines.
- Simultaneously, our refined language model (LLM) automatically extracted the CoD from the same 600 examples.
- A second trained nurse, acting as an adjudicator, independently reviewed both sets of results. This review ensured that the annotations adhered to the guidelines and that the primary CoD was accurately identified in each case.

Following the evaluation, we analyzed the results by determining the accuracy of primary CoD and additional identified causes from both the human annotator and the LLM per the adjudication. We then determined true positives, true negatives, false positives, and false negatives to compute relevant statistical metrics, allowing us to assess the accuracy and effectiveness of both human and automated CoD identification methods.

### Statistical metrics for model evaluation

For the transformer-based model evaluation, we calculated sensitivity, positive predictive value, and the F1-Score to evaluate model performance, and we computed micro averages for each to compute the average metric for a global measure of performance (All metric definitions are provided in the supplementary materials). We used Bootstrap to calculate the confidence interval by resampling the test set, calculating the required metrics for each resample, and using percentiles of these metrics to form the confidence interval. We also assessed the information density of online posts from each data source to determine their adequacy for reliable patient linkage and mortality information augmentation in healthcare systems.

For the LLM CoD information extraction module, we calculated the F-score, accuracy, precision, and recall for the primary CoD. However, for all potential CoD, due to the variation in the number of causes and the challenge of measuring performance using traditional NLP metrics, we asked the adjudicator to qualitatively assess the number of cases where the LLM correctly identified all the contributing CoD mentioned in the posts and to determine if the LLM and human annotators correctly identified the primary CoD. As such, we addressed the ambiguity of using a static output for each social media post. The adjudicator focused on whether the predicted CoD was accurate, regardless of whether it was explicitly mentioned in the post or inferred from the overall understanding of the post.

Phrases classified as “No CoD” indicated no specific medical CoD. These included “brief/sudden/extended/chronic illness,” “unexpected” or “sudden death/passing,” “natural causes,” “no mention” of cause, “none,” and “unknown/unspecified reasons/cause.” Posts containing only such phrases were categorized as “No CoD”. Correct identification of these cases by the language model counted as true negatives in the CoD identification process. This approach ensured that vague or non-medical descriptions were not misclassified as specific causes of death.

### Application of NLP and Final Data Collection

The final phase of our study involved compiling the extracted data into a comprehensive dataset ready for analysis. We applied a series of cleaning filters and NLP techniques to ensure that only documents with reliable mortality-related information were included. This thorough process resulted in a dataset, as detailed in Table 5 of the Results section. This dataset, enriched with mortality information from various sources, is poised to serve as a valuable resource for public health surveillance and future research efforts.

## Results

### Annotation inter-annotator agreement

Overall IAA with respect to GoFundMe achieved a 92.5% agreement rate in the final iteration while the IAA within Twitter data maintained an 85.7% agreement rate after 3 rounds of assessment. IAA achieved within data sourced from the obituary websites demonstrated strong overall agreement, with a 91.5% agreement rate after the third round of assessment.

### Information Density of Patient Identification in Social Media

Analysis of information density in online posts revealed varying levels of utility for patient linkage and mortality information augmentation in healthcare systems. Among the examined sources, three demonstrated high information density for annotated names, ranging from 87.81% to 97.81% (Table 1). These sources provided sufficient detail for reliable patient identification and mortality data enhancement, whereas X (formerly Twitter) had low information density for patient identification and was excluded from subsequent analysis.

**Table 1.** Information Density of Annotated Names Across Different Online Public Sources.

| Source Type | Names Annotated (%) |
| --- | --- |
| Twitter | 13.71 |
| Obituary | 96.48 |
| GoFundMe | 87.81 |
| Memorial Website | 97.81 |

### Extracting Mortality Information Results

Evaluated on the manually annotated test data, the RoBERTa model achieved the highest overall performance for extracting the targeted information, with a micro-averaged F1-score of 0.88 (95% CI, 0.86–0.90) (see Table 2). Confusion matrices are provided in the supplementary materials. This model outperformed others in all three tasks, achieving an F1-score of 0.85 (95% CI, 0.84-0.86) for Decedent Name, 0.89 (95% CI, 0.88-0.90) for Date of Death, and 0.94 (95% CI, 0.92-0.94) for Date of Birth. The ALBERT model attained an F1-score of 0.87 (95% CI, 0.86-0.89) for Date of Death, and F1-scores of 0.83 (95% CI, 0.82-0.86) for Decedent Name, and 0.91 (95% CI, 0.90-0.93) for Date of Birth. BERTweet achieved an F1-score of 0.90 (95% CI, 0.89-0.91) for Date of Birth, with scores of 0.82 (95% CI, 0.81-0.83) and 0.85 (95% CI, 0.84-0.86) for Decedent Name and Date of Death, respectively. BERT’s performance was marginally lower, with F1-scores of 0.81 (95% CI, 0.80-0.83), 0.84 (95% CI, 0.82-0.86), and 0.89 (95% CI, 0.88-0.90) for Decedent Name, Date of Death, and Date of Birth, respectively.

**Table 2:** Performance comparison of finetuned transformer models (on named entity recognition tasks (Decedent Name, Date of Death, and Date of Birth)

|  | RoBERTa |  |  | BERT |  |  | ALBERT |  |  | BERTweet |  |  |
| --- | --- | --- | --- | --- | --- | --- | --- | --- | --- | --- | --- | --- |
|  | Precision (PPV) | Recall (Sensitivity) | F1-score (95% CI) | Precision (PPV) | Recall (Sensitivity) | F1-score (95% CI) | Precision (PPV) | Recall (Sensitivity) | F1-score (95% CI) | Precision (PPV) | Recall (Sensitivity) | F1-score (95% CI) |
| **Decedent Name** | 0.86 | 0.84 | 0.85<br>(0.84-0.86) | 0.81 | 0.80 | 0.81<br>(0.80-0.83) | 0.84 | 0.82 | 0.83<br>(0.82-0.86) | 0.83 | 0.81 | 0.82<br>(0.81-0.83) |
| **Date of Death** | 0.87 | 0.91 | 0.89<br>(0.88-0.90) | 0.82 | 0.87 | 0.84<br>(0.82-0.86) | 0.86 | 0.88 | 0.87<br>(0.86-0.89) | 0.86 | 0.84 | 0.85<br>(0.84-0.86) |
| **Date of Birth** | 0.95 | 0.93 | 0.94<br>(0.92-0.94) | 0.90 | 0.89 | 0.89<br>(0.88-0.90) | 0.92 | 0.91 | 0.91<br>(0.90-0.93) | 0.91 | 0.89 | 0.90<br>(0.89-0.91) |
| **Micro Avg** | 0.88 | 0.88 | 0.88<br>(0.86-0.90) | 0.83 | 0.85 | 0.84<br>(0.82-0.86) | 0.85 | 0.87 | 0.86<br>(0.84-0.86) | 0.84 | 0.85 | 0.84<br>(0.82-0.86) |

The accuracy of the primary CoD identification and all CoD identification for both FSL-LLM and human identification are as follows: For GoFundMe, FSL-LLM achieved an accuracy of 95.9% for primary cause and 56.4% for all causes, while human accuracy was 97.9% for primary cause and 93.3% for all causes. For Obituary, FSL-LLM accuracy was 96.5% for primary and 96.0% for all causes, with human accuracy at 99.0% for primary causes and 98.5% for all causes. For Memorial websites, FSL-LLM accuracy was 98.0% for primary causes and 93.5% for all causes, whereas human accuracy was 99.5% for primary causes and 99.0% for all causes (Table 3).

**Table 3:**
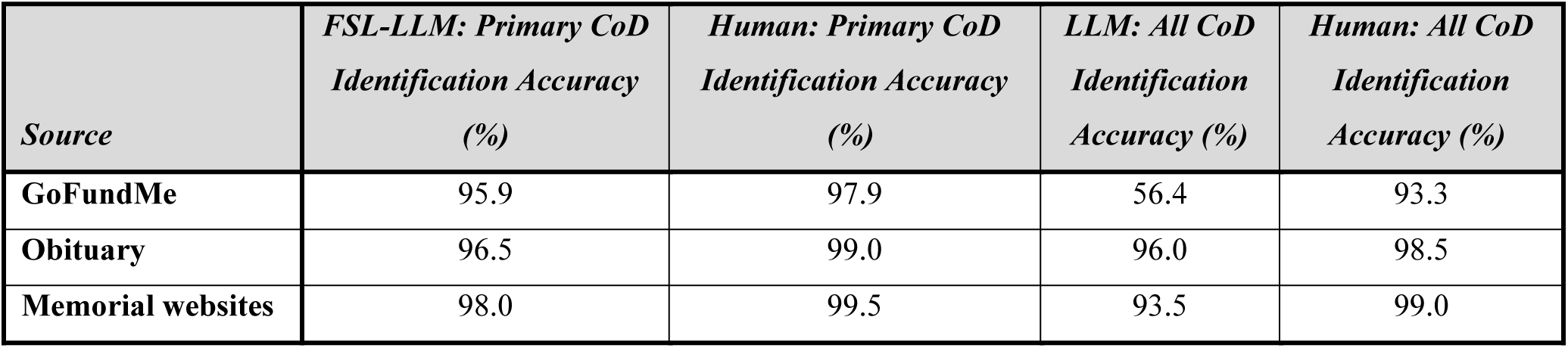
Accuracy of CoD Identification (FSL-LLM vs Human)

The precision, recall, and F-score for the LLM’s vs. human detection of the primary CoD were computed for each source. The metrics are presented below (see Table 4).

**Table 4:** Precision, Recall, And F-Score for FSL-LLM Vs human (Primary CoD)

| **Sources** | **FSL-LLM Precision** | **FSL-LLM Recall** | **FSL-LLM F1-Score** | **Human Precision** | **Human Recall** | **Human F1-Score** |
| --- | --- | --- | --- | --- | --- | --- |
| **GoFundMe** | 0.97 | 0.95 | 0.96 | 1.00 | 0.98 | 0.99 |
| **Obituary** | 0.61 | 1.00 | 0.76 | 1.00 | 0.82 | 0.90 |
| **Memorial Websites** | 0.94 | 0.98 | 0.96 | 1.00 | 0.98 | 0.99 |

**Table 5:** Number of Documents with mortality-related information identified from Each Source.

| Source | Total Documents |
| --- | --- |
| GoFundme | 23615 |
| Memorial website (Tribute archive; Ever loved) | 733754 |
| Obituaries.com | 7375229 |
| Total | 8,132,598 |

### Assessment of CoD Availability and Classification Error Analysis across Social Media Sources

As shown in Figure 3, social media sources varied significantly in the availability of CoD information. Obituaries had a very low density of CoD mentions (6%), while Everloved posts primarily contained a single potential CoD. GoFundMe was the richest source, with 43% of posts containing a single CoD and 50% containing multiple potential CoD mentions. However, not all mentioned causes of death or conditions pertained to the deceased subject of the social media post.

**Figure 3:**
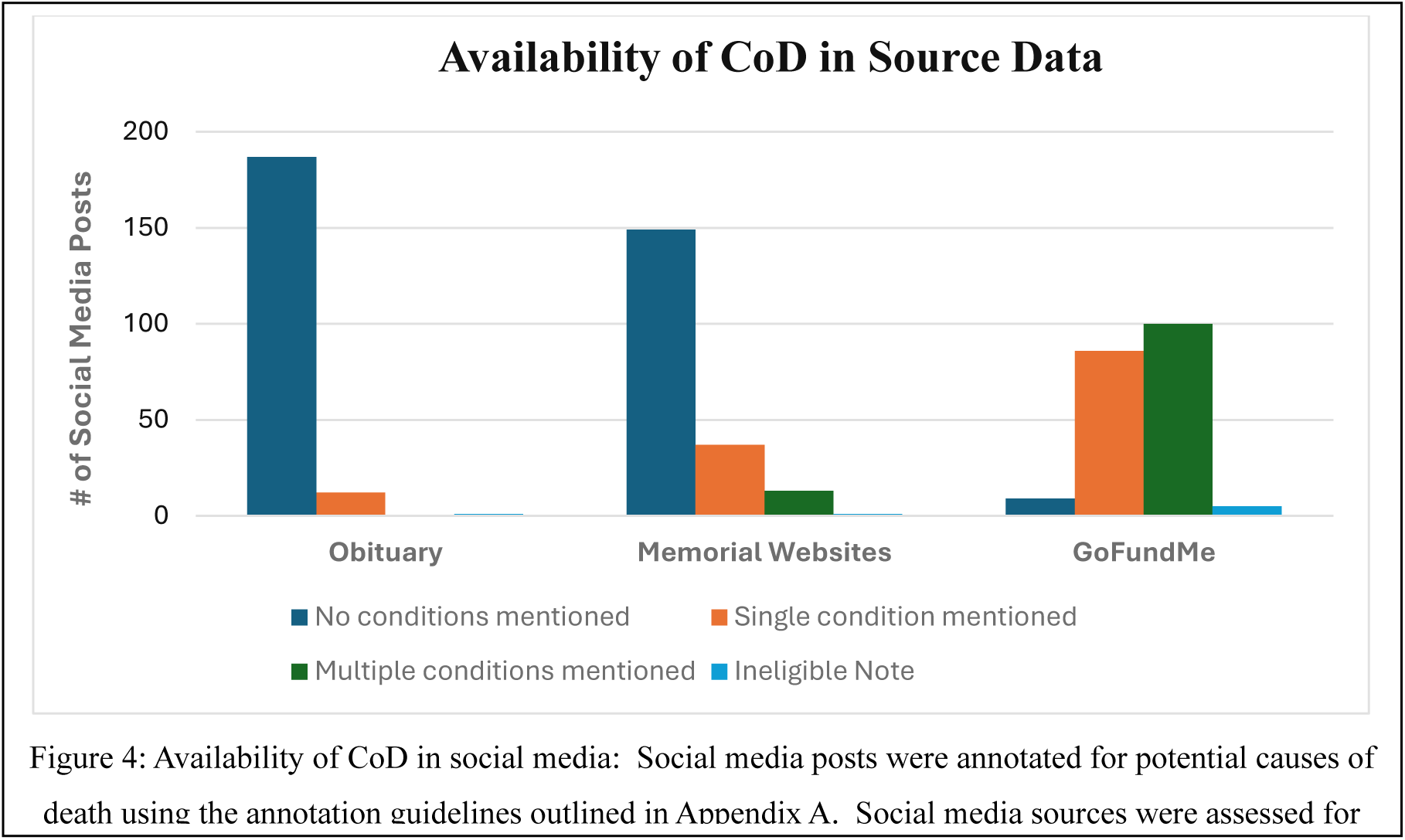
Availability of CoD within Social Media Posts.

The distribution and comparison of errors made by LLM and human annotators across the test dataset are illustrated in Figure 4. Each post may have multiple errors or error types. The analysis focuses on discrepancies in both primary and additional CoD annotations, providing a detailed breakdown of error types and frequencies. The errors include missed additional patient conditions, missed non-patient conditions, missed primary causes, incorrect CoD annotated, ineligible notes annotated, non-patient conditions attributed to the patient, and unclear annotation of no CoD.

**Figure 4.**
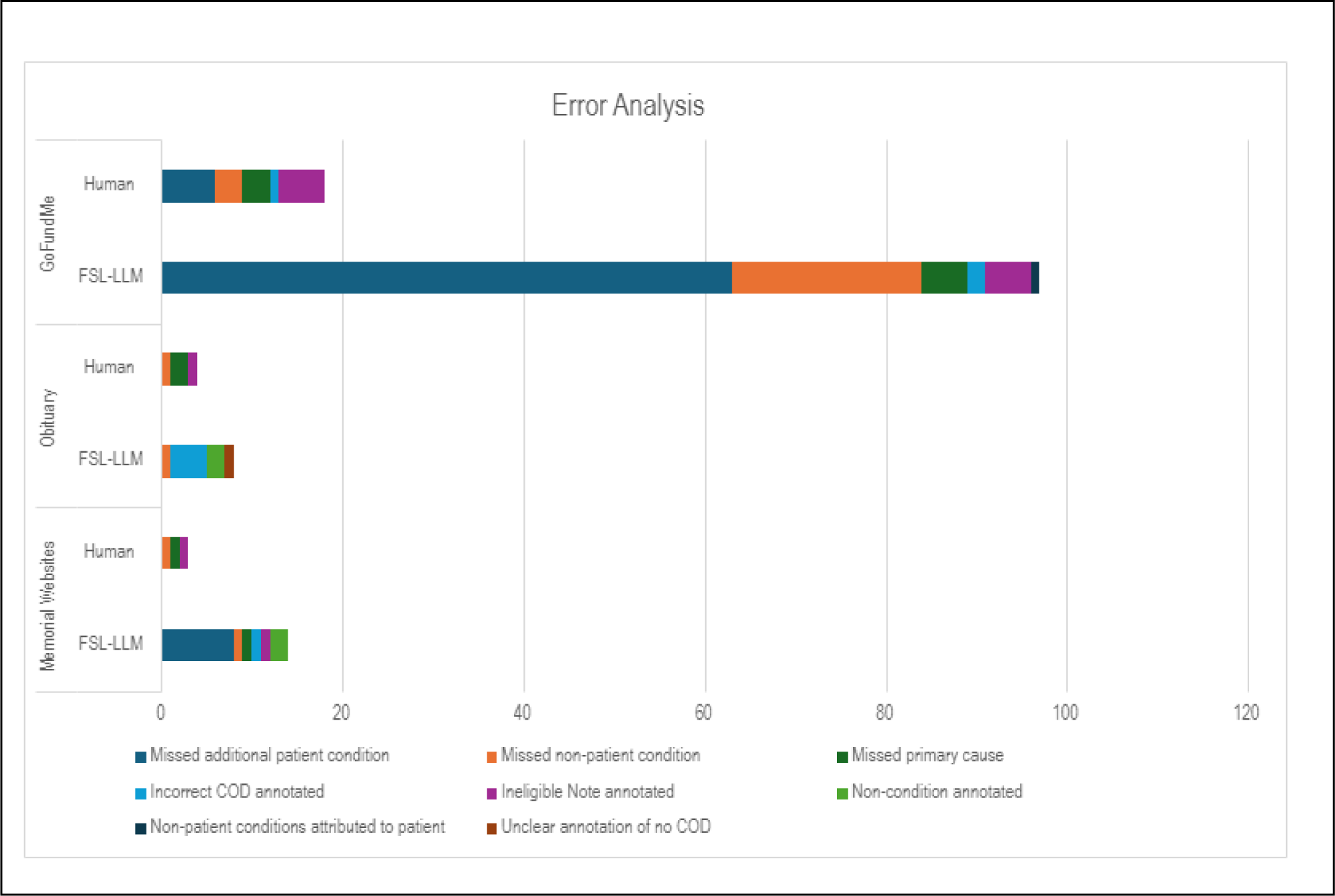
Types of errors between LLM vs. human annotation on the test dataset (errors per post)

The disparate information density across the data sources (see Figure 4) influenced the types of errors found within the annotations though the human annotator consistently had higher rates of agreement with the adjudicator than the computer annotations. Obituaries had a low density of CoD information and very low error rates. The most common error made by the FSL algorithm was in the annotation of a CoD that was not mentioned in the post (3.5%), whereas the human annotator missed a mentioned CoD in 1% of posts. For Memorial websites, both human and FSL-LLM annotations exhibited a small number of errors. FSL-LLM annotations missed mentions of medical conditions in 4.5% of posts and attributed primary causes incorrectly in only 2% of posts, whereas human annotators had an error rate of less than 1% for any category. For GoFundMe, which regularly mentions multiple patient and non-patient conditions, the FSL-LLM model has similar error rates to human annotation, except for “Missed non-patient condition” (10.5%) and “ Missed additional patient conditions” (31.5%) categories, indicating a performance gap compared to human annotations (1.5% and 3%, respectively) in identification of all potential medical conditions within the post though very low error rate in identification of the primary CoD.

### Final Collected Records

After applying the cleaning filters and NLP techniques, we successfully identified and extracted mortality-related information from a substantial number of documents across various sources. Table 5 below provides a summary of the total documents retained from each source.

## Discussion

We employed a novel approach to extract mortality data from online sources using transformer-based NLP models and few-shot learning with LLMs. Our analysis demonstrated the effectiveness of finetuned transformer-based NLP models in extracting mortality data from diverse online sources, showcasing their potential to enhance traditional data collection methods. We also developed a few-shot learning approach with LLMs to effectively identify primary CoD from online unstructured text data, achieving high agreement with human annotators. By leveraging publicly available online data, our approach has the potential to supplement conventional mortality databases, facilitating a more timely, comprehensive, and granular understanding of population-level mortality trends and risk factors.

Our study is consistent with other published papers that use social media data generally and obituary data specifically to improve the ability of health and healthcare research to accurately measure outcomes at the population level. For example, some studies have successfully used data from the Twitter platform to predict opioid overdose [39] and heart disease mortality [40], outperforming traditional demographic and health risk factors in predicting mortality. Additional studies have used GoFundMe data to identify disease categories in 89,645 medical crowdfunding campaigns [41] and to identify factors associated with cancer fundraising success [42]. An additional set of studies has used a range of techniques in online obituaries specifically, including for automated surveillance of cancer mortality trends [43], extraction of kinship data for genetic research [44], and reporting of drug overdose. [45]

Our study extends the existing literature by using transformer-based NLP models, which enhanced the extraction of key components of mortality data across public sources. Models such as RoBERTa, ALBERT, BERTweet, and BERT showed strong performance in handling unstructured data to extract decedent names (first and last), dates of birth, and dates of death, with RoBERTa achieving the highest micro-averaged F1-score of 0.88 (95% CI, 0.86-0.90).

For primary CoD identification, our FSL-LLM approach demonstrated high accuracy across all sources (GoFundMe: 95.9%, obituaries: 96.5%, memorial websites: 98.0%), approximating human annotator performance (97.9%, 99.0%, and 99.5% respectively). Detailed performance metrics revealed robust results for GoFundMe (precision=0.97, recall=0.95, F1=0.96) and memorial websites (precision=0.94, recall=0.98, F1=0.96). Obituaries achieved high accuracy, though the precision-recall pattern (precision=0.61, recall=1.0, F1=0.76) suggests potential for optimization in processing such data format. These findings demonstrate the model’s effectiveness while highlighting opportunities for source-specific improvements.

FSL-LLM demonstrated equivalent performance to human annotations for CoD identification across all sources; there remains room for further enhancement to identify potential contributing causes of death. The error analysis indicates that FSL-LLM exhibited higher error in categories such as “Missed non-patient condition” and “Missed additional patient condition,” whereas it exhibited very low rates of error in identifying primary CoD or appropriately classifying a note as having no specific CoD noted. This was primarily noted in GoFundMe data, as it was the only data source with significant posts containing more than one medical condition. Targeted improvements in the model’s ability to identify non-patient conditions and additional potential contributing causes are necessary to reduce these errors. The observed variation in error rates underscores the need for data-specific tuning to optimize model accuracy across different sources. To further enhance the FSL-LLM’s performance, focused finetuning on the identified error types and the integration of more diverse training datasets are recommended.

An additional finding was the low information density observed in the data from the X platform (Twitter at data collection time) relative to the other data sources allowing linkage to specific persons. The absence of reliable person identification in the data hinders reliable patient linkage, an essential element in the augmentation of mortality information and subsequent integration into the healthcare system. We therefore excluded Twitter data from the analysis, after the annotation phase.

Automated extraction of key mortality information from online sources has the potential to significantly improve traditional mortality databases, which often experience delays and incomplete data. This approach enables the timely collection of crucial details surrounding mortality such as decedent names, dates of birth, and dates of death, which could enable linkage to other healthcare data sources such as EHRs to facilitate clinical research. For instance, in studies monitoring medical product safety and effectiveness using insurance claims and EHR data such as in the FDA Sentinel system, mortality information from publicly available sources using approaches described here could allow investigators to study inferential questions regarding the impact of medical products on overall and cause-specific mortality. Integrating these methods with healthcare systems can improve reporting timeliness and completeness. [46]

### Limitations

Despite promising results, this study has several limitations. First, social media data may not fully represent all population segments due to usage and sharing biases. Second, although the NLP pipeline achieved high accuracy, the inherent ambiguity and scarcity of specific CoD mentions in the source data resulted in the underdetermination of some portions of the targeted information, as indicated by the human reference standard reviewers. Consequently, the NLP system may still misclassify some data points. Additionally, our reliance on keyword-based searches may miss relevant posts because of variations in language, slang, or indirect references to mortality, and the exclusion of non-English posts may limit generalizability. Moreover, social media posts may underreport stigmatized conditions such as HIV, suicide, or opioid-related deaths due to reporting bias, and the exclusion of non-users may further affect data representativeness. Finally, CoD identification from text remains challenging, often requiring an understanding of context and relationships between mentioned conditions. While the few-shot learning with the LLM algorithm performed well in identifying primary CoD, further work is needed to improve its ability to extract multiple contributing causes from individual posts. The use of a nurse adjudicator may also introduce correlated errors with the human-based reference, potentially providing a conservative estimate of model performance.

### Future Directions

At the population level, future research could focus on comparing CoD derived from online public data with those reported by official agencies. This comparison could help validate the accuracy and timeliness of online-sourced mortality information. If validated, such data could potentially provide near real-time insights into emerging mortality trends, particularly for rapidly spreading causes such as infectious diseases or environmental exposures.

The integration of online-sourced mortality data into existing surveillance systems would require careful validation against official records to ensure accuracy and reliability. This process would likely involve collaboration between researchers and public health agencies. Such collaborations could help develop protocols for effectively incorporating online data into public health surveillance and decision-making processes, potentially enhancing the speed and breadth of public health responses. Future research also should assess the plausibility of the CoD distribution from online sources by comparing it to sex- and age-adjusted national mortality statistics and investigating the potential underreporting of specific causes.

## Conclusion

We have demonstrated a promising application of advanced NLP techniques, including transformer-based models and few-shot learning with LLMs, to extract critical mortality information and identify causes of death from diverse online public data sources. The successful development of an NLP pipeline and the strong performance of the few-shot learning algorithm highlights the potential of these approaches to address limitations in traditional mortality databases and improve the timeliness, comprehensiveness, and granularity of mortality monitoring. However, the study acknowledges several limitations, such as potential biases in online data representation and challenges in extracting multiple contributing causes of death. Future research should focus on validating the usefulness of these methods in real-world settings, studying the correlation between online-derived causes of death and official records, and improving the integration of online data into public health surveillance systems. Addressing these challenges and opportunities will strengthen the application of advanced NLP techniques to online public data for enhancing mortality surveillance.

## Data Availability

All data produced in the present study are available upon reasonable request to the authors

## Code and Data Availability

The data supporting this study contain sensitive information and are therefore available upon request to ensure responsible use. The code can be downloaded at the following link:<u>Code</u>

## Ethics Approval and Consent to Participate

This study used publicly available online data and did not involve any interaction with human subjects. Data collection and analysis were conducted in accordance with ethical guidelines and fair use principles for research purposes.

## Author Contributions

**Conceptualization**: MA-G, RR, MEM, RJD, JJH-M. **Methodology**: MA-G, MLN-N, RJD, MEM. **Software**: MA-G, DW, MLN-N, MEM, MSK. **Validation**: JAD, JMW, MLN-N, RR, MFM. **Formal analysis**: MA-G, XW, AK. **Investigation**: MA-G, MLN-N, JMW. **Resources**: MEM, MM, RJD. **Data curation**: MFM, JAD, JMW, MLN-N, RR. **Writing – original draft**: MA-G, RJD, MEM, MLN-N, RR. **Writing – review & editing**: RR, RJD, MM, JJH-M, XW, AK. **Visualization**: MA-G, DW, MLN-N. **Supervision**: RR, MEM, RJD, JJH-M. **Project administration**: MLN-N. **Funding acquisition**: MEM, RJD, JJH-M, XW, AK.

